# Close contact infection dynamics over time: insights from a second large-scale social contact survey in Flanders, Belgium, in 2010-2011

**DOI:** 10.1101/2020.09.30.20204891

**Authors:** Thang Van Hoang, Pietro Coletti, Yimer Wasihun Kiffe, Kim Van Kerckhove, Sarah Vercruysse, Lander Willem, Philippe Beutels, Niel Hens

## Abstract

**Background:** In 2010-2011, we conducted a social contact survey in Flanders, Belgium, aimed at improving and extending the design of the first social contact survey conducted in Belgium in 2006. This second social contact survey aimed to enable, for the first time, the estimation of social mixing patterns for an age range of 0 to 99 years and the investigation of whether contact rates remain stable over this 5-year time period.

**Methods:** Different data mining techniques are used to explore the data, and the age-specific number of social contacts and the age-specific contact rates are modelled using a GAMLSS model. We compare different matrices using assortativeness measures. The relative change in the basic reproduction number (*R*_0_) and the ratio of relative incidences with 95% bootstrap confidence intervals (BCI) are employed to investigate and quantify the impact on epidemic spread due to differences in gender, day of the week, holiday vs. regular periods and changes in mixing patterns over the 5-year time gap between the 2006 and 2010-2011 surveys. Finally, we compare the fit of the contact matrices in 2006 and 2010-2011 to Varicella serological data.

**Results:** All estimated contact patterns featured strong homophily in age and gender, especially for small children and adolescents. A 30% (95% BCI [17%; 37%] ) and 29% (95% BCI [14%; 40%] ) reduction in *R*_0_ was observed for weekend versus weekdays and for holiday versus regular periods, respectively. Significantly more interactions between people aged 60+ years and their grandchildren were observed on holiday and weekend days than on regular weekdays. Comparing contact patterns using different methods did not show any substantial differences over the 5-year time period under study.

**Conclusions:** The second social contact survey in Flanders, Belgium, endorses the findings of its 2006 predecessor and adds important information on the social mixing patterns of people older than 60 years of age. Based on this analysis, the mixing patterns of people older than 60 years exhibit considerable heterogeneity, and overall, the comparison of the two surveys shows that social contact rates can be assumed stable in Flanders over a time span of 5 years.

## Introduction

Infectious diseases and, more specifically, airborne infections can be transmitted between hosts via close contact interactions; therefore, quantifying such interactions provides important information for properly modelling infectious disease transmission. In recent years, we have witnessed a paradigm shift with respect to this: whereas at the start of this century, mathematical models relied on simplifying assumptions such as homogeneous mixing or on using mathematically convenient “Who Acquires Infection From Whom” constructs [1], a vast number of studies now rely on the use of social contact data [2, 3, 4, 5, 6, 7, 8].

The literature on social contact surveys has shown how human interactions are heterogeneous in nature and present a large degree of homophily in terms of age [9, 10] and gender [11]. The information coming from social contact surveys is therefore usually summarized in what is called the social contact matrix, quantifying the average number of contacts made between individuals within and between given age classes. Using the *social contact hypothesis* [2], i.e. assuming that transmission rates are proportional to social contact rates, these data-driven mixing patterns have been implemented into models of infectious disease transmission showing good correspondence to (sero)prevalence data; see, e.g., [4, 9, 12].

Social contact survey data allow for an exploration of contact rate patterns stratified by age, gender, and location, which helps to better describe the structure of the transmission network [13, 14]. However, a systematic review by Hoang et al. (2019) [10] showed that half of the social contact surveys before 2019 used convenience sampling, while quite a few surveys were conducted in specific settings, e.g., schools or universities, and/or focus on specific target groups; thus, it is impossible to extrapolate the results to an entire population. Even in population-based social contact surveys with representative samples, two problems might still exist: the sample does not cover all age ranges of the population, or the number of elderly participants is insufficient for investigating mixing patterns of these people. Indeed, no study reported the contact rates of people up to 99 years old.

Particular attention has been devoted to behavioural changes with respect to individual health status (e.g., being ill [6, 15, 16, 17]), weather conditions [5] or day of the week (weekday or weekend in holiday/non-holiday or regular periods [9, 18, 19, 20, 21, 22]) - hereafter referred to as *microscopic* time settings, and how these affect disease dynamics [6, 8, 19, 23, 24].

The use of social contact data to inform modelling has become so prominent in recent works that it has also been applied to settings for which social contact studies are not available, leading to the question of how social contact matrices should be projected onto other geographical areas and in time [7, 25, 26, 27]. However, to the best of our knowledge, there has been no empirical assessment of whether mixing patterns change over longer time periods (e.g., years) within a particular population and how this should be taken into account when projecting social contact matrices. We will refer to these as *macroscopic* time changes to mark the difference with *microscopic* time changes.

A first population-based social contact survey in Belgium was conducted in 2006, and its results were reported in [9, 19, 28], in which the impact of *microscopic* time changes on the contact mixing pattern was investigated, although this study was not designed for doing so. A second population-based survey in Belgium was conducted 5 years later in 2010-2011. This survey was conceived as an improvement over the 2006 survey, with a larger sample size covering a wider age range of participants and a better distribution of surveyed participants over four different time settings (weekday/weekend days in regular/holiday periods).

In this work, we aim to describe and analyse the Flemish social contact survey from 2010-2011 by accounting for the mixing patterns of people 0-99 years of age, with a special focus on elderly people. We study both the impact of *microscopic* and *macroscopic* time changes on contact patterns, and we assess whether the contact rates remain stable over 5 years timespan.

## Data

### Social contact survey in 2006

This survey was part of the POLYMOD project, in which social contact surveys were conducted in 8 European countries in 2005-2006 [9]. In the social contact survey conducted in Belgium in 2006, a total of 750 participants were recruited by random digit dialing on land lines. The survey sample covered all three regions in Belgium; i.e. the Flemish, Walloon and Brussels-Capital regions, with quota sampling by age, gender, and region, making it representative for the whole Belgian population. Each participant was asked to fill in a background questionnaire and a paper diary in which they record their contacts over 2 days: one randomly assigned weekday and one randomly assigned weekend day. Two types of contacts were defined: (1) two-way conversations during which at least three words were spoken and (2) contacts that involved skin-to-skin touching. Information recorded in the diary included the gender and the exact age or presumed age interval of each contacted person over the entire day. Contact features included frequency, location and duration. If participants established more than 20 professional contacts per day, then they only had to provide an estimated number of professional contacts and the age interval(s) with whom they interacted most. Contact information (e.g., contact age or contact duration) was then imputed for such contacts. More details can be found in [28]. We will refer to those contacts as additional professional contacts.

### Social contact survey in 2010-2011

This survey was conducted between September 2010 and February 2011 in the Flemish region in Belgium using an adapted version of the diaries used in the first Belgian survey in 2006. Three different types of diaries were designed to adapt to the age of participants: one for children (less than 13 years old), which was completed by a proxy, e.g., parents or school teachers; one for people aged 13-60 years and one for people aged 60+ years, which could also be filled out by a proxy. A total of 1,774 participants were recruited by random digit dialing on mobile phones and landlines, with quota sampling by age, gender and geographical location. The contact definitions were the same as those used in the 2006 survey. Participants were asked to complete a background survey and record their social contacts in a paper diary during one randomly assigned day. Information on additional professional contacts was imputed the same way as done for 2006 data. Compared with the 2006 survey, the 2010-2011 survey explored more features that might influence the number of contacts recorded: the health conditions of participants, time use, distance from home, animal ownership and touching. To date, the impact of animal ownership and touching on social contacts has been investigated [29],so has the impact of weather on social contacts [5]. Of particular focus were people aged 60 years and above; i.e., participants up to 99 years of age were recruited, and information about contact frequency with children and grandchildren and residence size for elderly people living in nursing/elderly homes was recorded.

The design of the 2010-2011 survey is similar to that of 2006, with the difference being that in 2010-2011, participants reported information for only one day, whereas in 2006, information was collected for two days. Since participants have been shown to be influenced by fatigue in reporting on multiple days [10, 22], only data on the first day of the 2006 survey will be used for comparison with the 2010-2011 survey in this work. Additionally, we extracted the 511 participants recruited in Flanders in the 2006 survey to be in line with the surveyed regions in the 2010-2011 survey. In the 2010-2011 survey, 15 cases were removed since the diaries were unreliable (many answers left blank, incoherent answers, etc.). We also excluded 46 people living in an elderly/nursing home and explored the contact patterns of these people separately; in addition, 6 people aged more than 90 years were removed to avoid problems related to data sparsity. As a result, the final sample for the analysis of the 2010-2011 survey is 1,707 participants. We defined four microscopic time settings: regular weekdays, regular weekends, holiday weekends, and holiday weekdays. Holiday periods include both public holidays and weekends inside or adjacent to these holidays. More details on the number of participants by age and microscopic time in both surveys can be found in SA1 Table S1. The datasets of both surveys are available online within the *social contact data sharing* initiative [30] and the SOCRATES platform [14].

## Methodology

We start with a descriptive analysis to explore the socio-demographic characteristics of survey participants and features of their reported contacts for the contact survey in 2010-2011. Subsequently, data mining techniques are used to explore associations among variables of interest and contact profiles of survey participants. We then investigate the factors associated with the number of contacts, differences in gender in mixing patterns and the impact of holidays and weekends as a proxy for the impact of school closure on disease transmission. We end with the comparison between the contact surveys from 2006 and 2010-2011 using different measures.

### Data mining techniques

We use two unsupervised learning methods: association rules and clustering. Association rules are used to assess the possible associations pertaining to contact features, e.g., type of contact (close or non-close), duration and frequency of contacts, …, using support, confidence and lift values as measures of interestingness [19, 31] (see SA2 for additional information). Rules are considered of interest only when the support value exceeds 1%, equivalent to at least 3142 contacts involved in constructing the rules. The threshold for the confidence is 70%, and rules with greater lift indicate stronger association. In addition to association rules, we investigate contact profiles using a clustering method. The contact profiles are defined by (1) the number of contacts per survey participant in six different locations (home, work, school, leisure, transport and other), (2) characteristics of participants (age and gender) and (3) time indicators (weekday/weekend and regular/holiday period). Clustering is implemented using the *daisy* function in the R package “Cluster” [32] and using the *Gower distance*, which allows for mixed types of variables. We visualize the clusters by projecting them into a low-dimensional space using a dimension reduction technique known as the *t-distributed stochastic neighbor embedding* [33]. Contacts reported at different locations are attributed to just one location using the following hierarchy: (1) contacts at home, (2) contacts at work, (3) contacts at school, (4) contacts during leisure time, (5) contacts in other locations and (6) contacts in transportation.

### Modelling the number of contacts

#### Degree distribution

We consider both physical and non-physical contacts, including additional professional contacts reported by participants. We model the number of contacts using a weighted negative binomial regression model to account for over-dispersion. Socio-demographic characteristics, the health status of participants and microscopic time settings (weekdays/weekends and regular/holiday period) are included as possible determinants (descriptive statistics see SA1). In addition, diary weights computed from age and household size are used to account for under-/over-sampling over participant features [9, 28]. We perform variable selection using a random forest analysis [31] and the likelihood ratio test (LRT). Interactions between age and microscopic time settings are retained, as they are the two most significant determinants of the number of contacts reported in the literature [10].

#### Estimating age-specific contact rates

We define the age-specific number of contacts *y*_*ijr*_ as the number of contacts made by the *r*^*th*^ participant in age class *i* with people in age class *j* per day (*i, j* = 1, *…, J*; *r* = 1, *…, n*_*i*_), where *J* is the number of age classes, and *n*_*i*_ is the number of participants in age class *i*.

The age-specific number of contacts *y*_*ijr*_ is assumed to follow a negative binomial distribution to account for over-dispersion [9]. This distribution is defined as *y*_*ijr*_|***x*** ∼ *NB*(*m*_*ij*_, *κ*_*ij*_) for a vector of covariates ***x***, in our case the age of the participant *x*_1*i*_ and the age of the contact *x*_2*j*_. The mean and variance of this distribution are defined as *m*_*ij*_ and 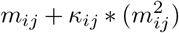 respectively, where *κ*_*ij*_ is the over-dispersion parameter. To model the age-specific number of contacts, we apply generalized additive models for location, scale and shape (GAMLSS). This allows for modelling both the mean and variance (over-dispersion) parameters of the negative binomial distribution over participants’ age *x*_1*i*_ and contacts’ age *x*_2*j*_. We refer to SA3 and [34] for details about the GAMLSS. When estimating the social contact matrix *C*, the reciprocal nature of making contact needs to be taken into account, as *m*_*ij*_*N*_*i*_ = *m*_*ji*_*N*_*j*_, where *N*_*i*_ is the population size in age class *i* (obtained from demographic data) [35]. Based on *m*_*ij*_ and *N*_*i*_, the reciprocal contact rates *c*_*ij*_ can be obtained by 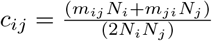.

For all quantities of interest, introduced in this and following subsections, we use a non-parametric bootstrap of participants, to obtain 95% percentile bootstrap confidence intervals (BCIs) [36].

#### Measures of comparison between different mixing patterns

We use four different measures of comparison: two for measuring assortativeness, the relative change in *R*_0_ and the relative incidence (RI). We measure the assortativeness of contacts by age using 2 different indices. The first index is Gupta’s Q [37], which ranges from 0 (= homogeneous mixing) to 1 (= completely assortative mixing). The second index is 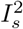, as proposed in [38] ranging from 0 (= perfect assortativity) to 1 (= homogeneous mixing).

The third measure is based on the basic reproduction number *R*_0_. *R*_0_ is given by the dominant eigenvalue of the next generation matrix ***G***. Assuming the age-specific transmission rates *β*(*i, j*) are proportional to the age-specific social contact rates *c*(*i, j*) (also known as the social contact hypothesis [2]), the ratio of dominant eigenvalues of the next generation matrix yields the relative change in basic reproduction number using different mixing patterns. Lastly, the ratio of relative incidences (RRI) is used for comparison. The expected age-specific RI in the population during the exponential phase is given by the leading right eigenvector of the next-generation matrix [39].

For more details, we refer to SA3.

#### Investigating gender differences in mixing patterns

To gain insights into possibly different mixing behaviour between males and females, we estimate the age-specific average number of contacts using the GAMLSS approach (as previously introduced) for all four combinations of gender interactions (male-male, female-female, male-female and female-male). We use assortativeness measures (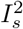 and Q indices), and the RRI to study differences between matrices.

#### Investigating the impact of school closure on disease transmission

We estimate the impact of school closure based on social contact data using contact rates from holidays and weekend days as a proxy and compare them with contact rates from regular weekdays. We use the changes in *R*_0_ and the RRI to quantify these differences.

### Comparing the contact surveys in 2006 and 2010-2011

The designs of the contact surveys in 2006 and 2010-2011 are similar (see Section Data). However, the 2006 survey did not cover age groups up until 90 years of age or sufficiently cover all four microscopic time settings (see SA1 Table S1). Therefore, for the comparison of contact matrices between 2006 and 2010-2011, we only use participants less than 65 years old and merged weekend-regular and weekend-holiday into one “weekend” category to overcome the data sparsity problem.

#### Degree distribution of contacts from aggregated data

To investigate potential determinants for the number of contacts, we combine data from 2006 and 2010-2011 using a survey indicator variable as an additional determinant. We first apply a random forest analysis [31] to identify main predictors, after which we model the aggregated number of contacts via a GAMLSS model assuming a negative binomial distribution for the response variable. Model selection is performed using the likelihood ratio test for mean and dispersion parameters.

#### Comparing contact matrices

We first visually compare contact matrices in 2006 and 2010-2011 for the different microscopic time settings. Both assortativeness indices, the change in *R*_0_ and the *RRI* are used to compare mixing patterns. Furthermore, we use the ratio of transmission rates that allows for the direct comparison of contact rates between contact matrices (a cell-wise comparison). Lastly, we applied the methods outlined in [4] to use social contact matrices to fit VZV serological data from Belgium based on the social contact hypothesis (i.e., constant proportionality, [2]), with both contact data sets separately and compare the results.

## Results

### Socio-demographic characteristics of participants

A total of 1,707 participants (46% males and 54% females) are used for the analysis of the 2010-2011 dataset, of whom 1,011 reported their contacts on regular weekdays, 257 on regular weekend days, 286 on weekdays during public holidays and 151 on weekend days during or adjacent to public holidays (2 cases did not indicate the date). The average participant age was 38 years, and participants younger than 18 years of age accounted for 22% of the sample. The average household size was 3, ranging from 1 to 11; participants with household sizes of 2-4 accounted for nearly 75% of the sample size, while only 13% of participants lived in households with more than 4 residents. Approximately 21% of the participants were still students, 48% had a job, 13% were retired and approximately 11% were at home or unemployed. Nearly two-thirds of working participants were office clerks, 19% were manual workers, and only 6% were self-employed.

### Daily number of contacts and contact features

A total of 31,423 contacts including 8,527 imputed professional contacts were recorded by the 1,707 participants: the highest number of contacts reported by one participant was 364 (mostly professional contacts), and the lowest was zero (15 cases). The median number of reported contacts were 12 (interquartile range (IQR):[6; 21]). Participants reported an average of 18.4 contacts per day (SD=24.3), skewed in distribution. This reduces to 13.4 (SD=10.8) when professional contacts are excluded. By adjusting for the age and household size of the Flemish population and type of day (weekdays/weekend days), the average number of contacts equals 17.2 (12.0 when excluding professional contacts).

Nearly half the number of contacts involved touching (with missing information in 345 cases). More than 10% of all contacts were with household members. Daily contacts accounted for nearly one-third of the total number of contacts, while only 10% were first-time contacts. Short contacts (less than 5 minutes) made up approximately 15% of the total number of contacts; long contacts (longer than 1 hour) constituted nearly half of the total number of contacts. Nearly two-thirds of all reported contacts were made at home, work and school, while contacts at multiple locations accounted for only 6% of all contacts.

### Data mining techniques

The association rules with the highest lift value are presented in Table S1 in SA2. Seventy-four percent of daily contacts lasting longer than 4 hours involved skin-to-skin touching. In contrast, 81% of the contacts lasting less than 5 minutes with non-household members were usually non-physical contacts. Contacts with household members are the most influential factor in determining whether contacts occur on a daily basis. Contacts lasting longer than 4 hours, occurring on weekdays in a regular period, tend to occur on a daily basis (71%).

In the clustering analysis, the largest silhouette width is obtained for six clusters (Figure S1 in SA2). The cluster sizes ranged from 151 participants (cluster 5) to 443 participants (cluster 1). All clusters present a strong connection with the microscopic time settings, including participants from only weekdays/weekends or regular days/holidays (Table S2 in SA2). Some clusters are easy to interpret when looking at the cluster members’ features. Cluster 2, for example, is composed of participants whose average age is 9 years, with a large number of contacts at school, i.e., school-aged children. Cluster 4 includes participants with an average age of 39 years and a large number of contacts at work, i.e., working-age adults. Other clusters present less specific contact patterns but still exhibit a strong connection with microscopic time settings. Specifically, cluster 1 includes participants who have a low number of contacts in all locations on regular weekdays. Participants in this cluster have the highest average age (51 years) and can be interpreted as being socially *non-active*. Cluster 3 includes participants surveyed on the weekend and regular period, with few contacts at work and school but the highest number of contacts during leisure activities and at “other” locations. Cluster 5 contains participants surveyed in the weekend and holiday period, with no contacts at school, few contacts at work and most contacts at home and in “other” locations. Cluster 6 consists of participants surveyed in the weekday and holiday period, with an average of 6 contacts at work, a very low number of contacts during leisure activities and transportation (see SA2 Table S2 and Figure S2).

### Degree distribution for the social contact survey in 2010-2011

Figure 1 shows the results of the weighted negative binomial model for the number of contacts. Several sociodemographic indicators have a significant effect on the number of contacts: age, household size, place of living and the use of public transportation. It is noted that occupation and education have been excluded from the model, as they strongly correlate with age, and that for participants younger than 13 years, the mother’s educational level is used instead of that of the participant. Other variables (gender, animal ownership, health states regarding self-care and pain) were also excluded after model selection was performed (see Table S1 and Figure S1). The interactions between age and microscopic time indicators are highly significant. In Figure 1, we compare the number of contacts among age groups in each time setting: for weekdays in the regular period, participants older than 5 years of age have a higher number of contacts than children aged 0–5 years, except the 15–25 age group, and people older than 60 years who have the lowest number of contacts. For holiday weekdays, people between 40 and 50 years of age have the highest number of contacts, while other age groups show no difference in the number of contacts relative to the youngest age group (0–5 years). The number of contacts of children aged 0–5 decreases by 25%, 46% and 54% with respect to regular weekdays during holiday weekdays, regular weekends and holiday weekends, respectively. During regular periods, people aged 75–90 have almost double the contacts during weekends with respect to weekdays. A higher number of contacts is observed in participants living in larger households and those using public transportation. The number of contacts is 41% lower in those who felt ill on the survey day. The reported health indicators show that feeling anxiety and having problems in carrying out daily activities have a negative effect on the number of contacts, reducing them by 18% and 35%, respectively.

**Figure 1:**
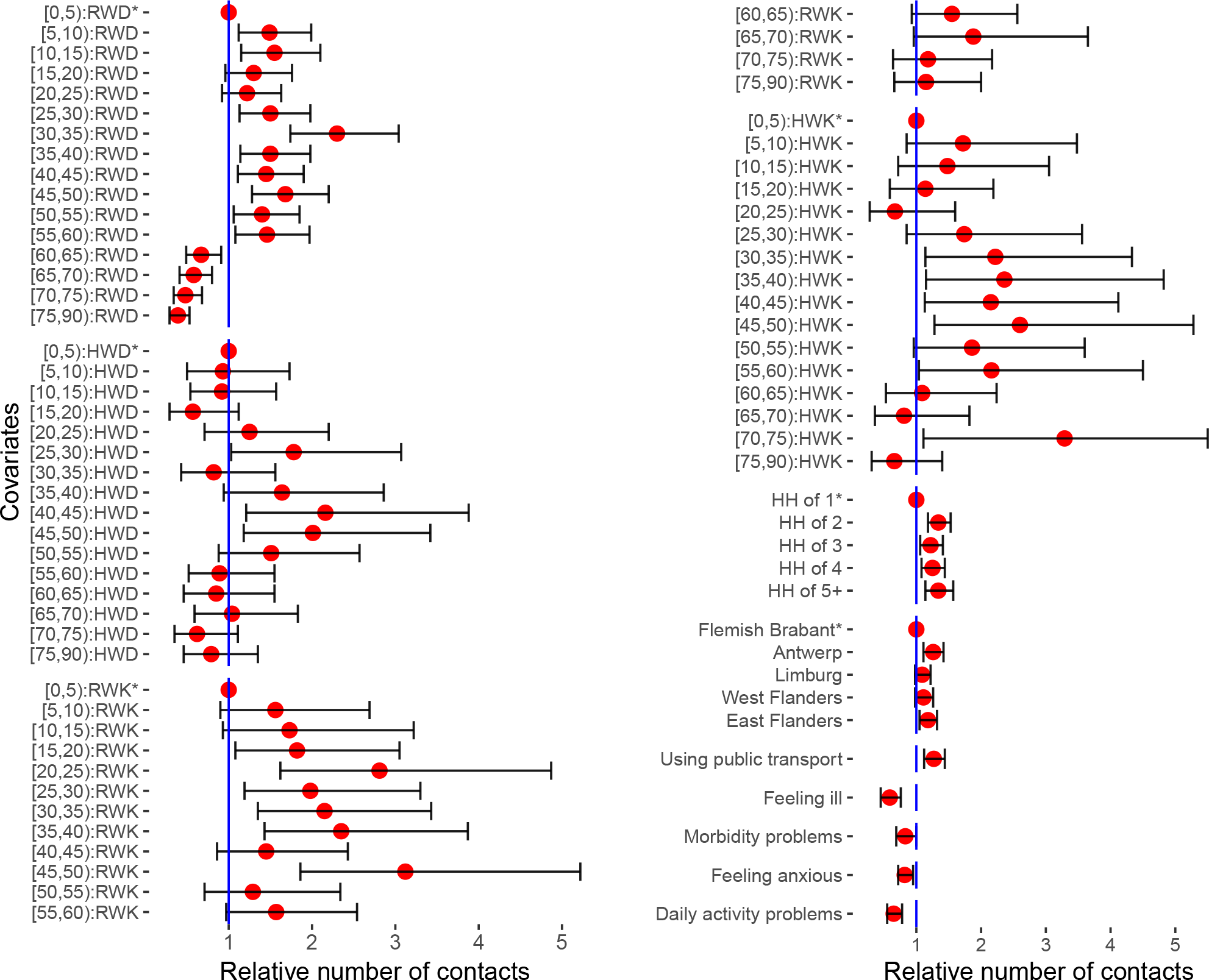
The results of a weighted negative binomial regression model for the total number of contacts in Flanders, Belgium, in 2010–2011. Relative number of contacts (red dot) and 95% confidence intervals (interval) based on a weighted negative binomial regression model for the total number of contacts in Flanders, Belgium, in 2010–2011 (n = 1,705, excluding 2 cases with missing information on the survey date). RWD, HWD, RWK and HWK stand for regular weekdays, holiday weekdays, regular weekends and holiday weekends, respectively. Stars (*) indicate reference groups for covariates with more than 2 categories.

### Contacts of children, working people and elderly people

Some variables were only present in diaries for children (less than 13 years old), adults or people older than 60 years of age, so these variables were not included in the previous model (Figure 1). Children attending preschools or schools have more contacts than children at childcare outside home and young children who are kept at home (P*<*0.001, Kruskal-Wallis test). It is also observed that there is no difference in the number of school contacts among different class sizes (P=0.40, Kruskal-Wallis test). Children less than 3 years old have nearly 40% of their contacts at home. School contacts make up 44% of the number of contacts for participants younger than 18 years old, and this figure increases to 60% on weekdays and during regular periods. People with a job have a much higher number of contacts than those who are retired or currently unemployed/job seeking. Working people have most contacts at work (63% on a random day and 71% on regular weekdays). When considering occupation, the number of contacts of office clerks is observed to be significant higher than those of people with other occupations.

Modelling the number of contacts for participants older than 60 years who are not living in an elderly/nursing home (SA3 Table S2) shows that drinking status has no significant effect on the number of contacts and neither does having children or grandchildren. Smokers have fewer contacts than those who are non-smokers or used to smoke. Elderly people who experience problems in performing their daily activities have fewer contacts than those who do not experience problems (relative number of contacts (RNC): 0.64; 95% CI [0.49; 0.84]). The effect of microscopic time settings is significant: more contacts were observed in weekend-regular periods than in weekday-regular periods. The majority of elderly people who have children and/or grandchildren reported having contacts with their children and grandchildren a few times per week or month. Figure 2 describes the social interaction of people aged 60+ years with other age groups. People aged 61–79 years have the highest number of contacts with age group [40, 60), which may describe the mixing pattern of people from 2 generations. Interaction between people aged 60+ years and young children/teenagers is significantly higher on holiday-weekdays and weekend days than on regular-weekdays.

**Figure 2:**
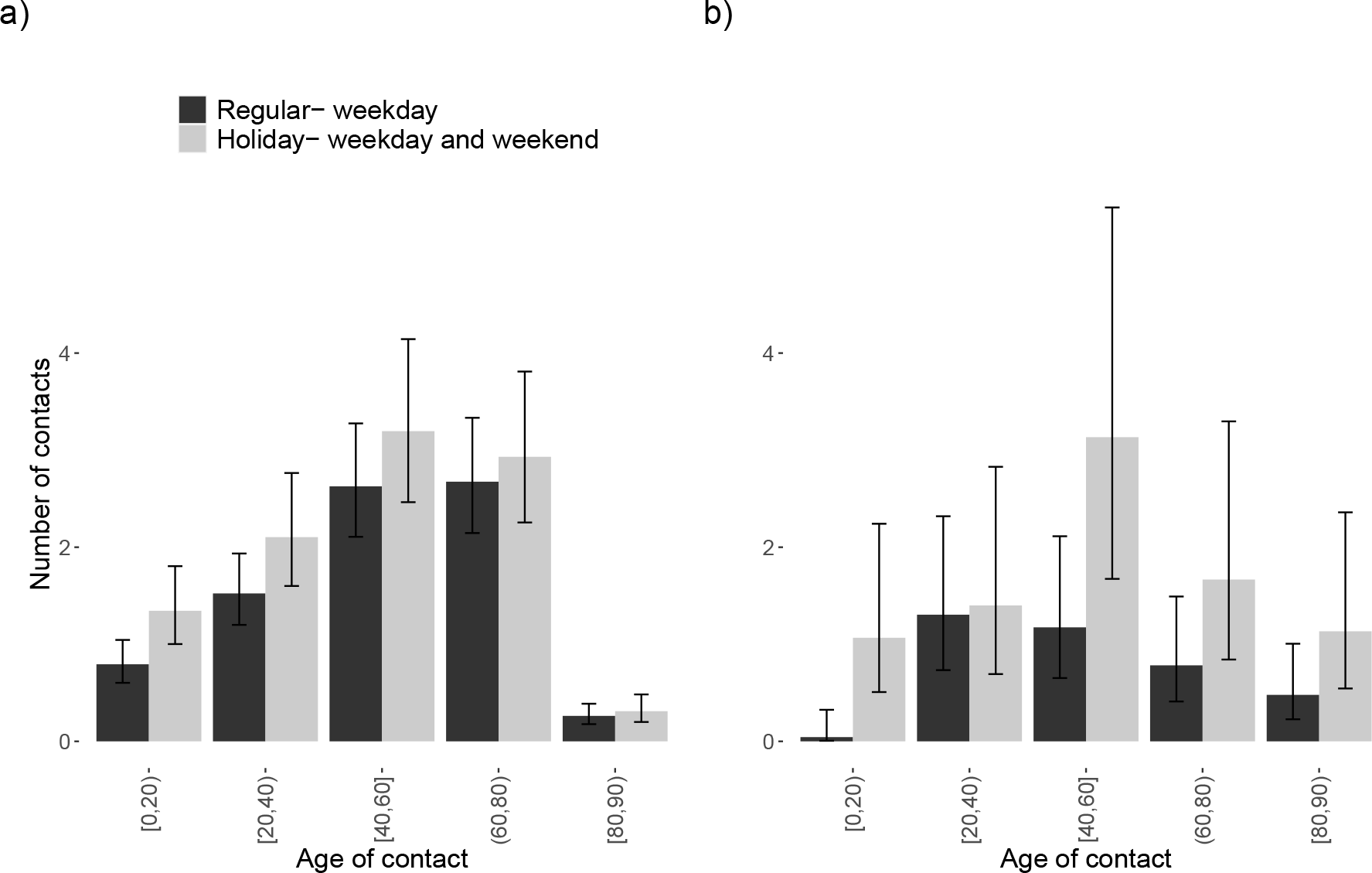
Age-specific number of contacts of older people for the 2010-2011 survey. (a): Age-specific number of contacts of people aged 61–79 years. (b): Age-specific number of contacts of people aged 80+ years.

### People living in an elderly/nursing home

Forty-six people reported living in a nursing/elderly home, with ages ranging from 79 to 99 years. Most of them have health problems: some problems or not being able to perform their daily activities (96%), some problems walking (67%) or staying in bed all the time (15%); some problems with self-care (46%) or not being able to care for themselves (43%); and experiencing mild to serious pain (85%) and anxiety (48%). These people reported 13.7 contacts on average, significantly higher than those aged 60+ years and living at home (P<0.0.001, Mann-Whitney test). No statistically significant difference in the number of contacts for people in elderly/nursing homes was found with respect to the residence size (P=0.47, Kruskal-Wallis test for 3 groups of residence sizes: <50, 50–100 and 100+). We compared people living in an elderly/nursing home with people aged 60+ years living at home with respect to their social interaction with other age groups (see SA3 Figure S2): almost no interaction with young children and teenagers is observed for people living in an elderly/nursing home, while this interaction is more observed for people aged 60+ years living at home.

### Overall contact patterns

The contact patterns by age group were summarized in a contact matrix displaying ages from 0 to 90 years, whose elements represent the contact rate between an individual in a given age group and an individual in another age group in the Flemish population. The resulting contact matrix shown in Figure 3 is described by the pronounced main diagonal indicating contacts with individuals in the same age group, e.g., at home, at school and at work, and the 2 less-pronounced sub-diagonals representing contacts between generations, e.g., children and their parents.

**Figure 3:**
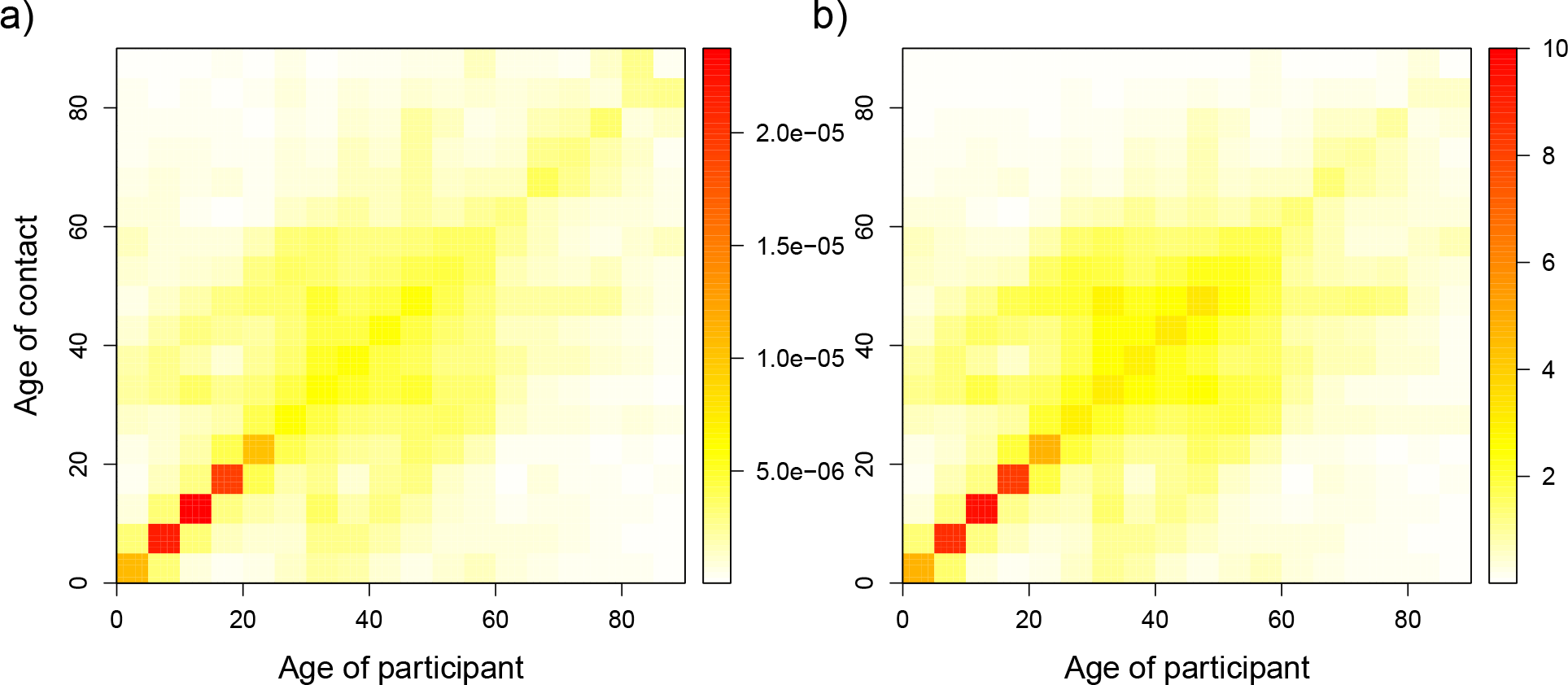
Estimated age-specific contact rates and average number of contacts for the 2010-2011 survey. (a): Estimated symmetric age-specific contact rates. (b): Age-specific average number of contacts. The color scale indicates the contact rates from low (white) to high (red).

### Gender differences in mixing patterns

Figure 4 shows the age- and gender-specific average number of contacts. The assortative mixing pattern characterized by the main diagonal is still observed for all interactions. The measures of assortativeness for all ages provide no differences for same gender vs. different genders, with overlapping 95% BCI of 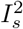 indices (0.39 [0.35; 0.45] and 0.42 [0.40; 0.45] vs 0.48 [0.43; 0.54] and 0.44 [0.42; 0.47], for male-male and female-female vs. male-female and female-male, respectively). Focusing on people aged less than 30 years, we noticed that the assortative mixing pattern is more pronounced in male-to-male and female-to-female contacts. Specifically, for contacts made between people in the same age groups, the average number of male-to-male contacts and female-to-female contacts are 3.9 (BCI [3.5; 5.0]) and 4.2 (BCI [3.7; 4.9]), respectively. These figures were reduced to 2.3 (BCI [2.0; 2.8]) for male-to-female contacts and 2.6 (BCI [2.3; 3.1]) for female-to-male contacts. The inter-generational mixing pattern (mostly parent-child), marked by the two sub-diagonals, is similar for females and males. The relative incidences for both genders also follow a similar pattern, with peaks at approximately 15 years of age and between 40 and 45 years of age, and no difference is found in the overall RI of males compared to that of females (see Figure S3 in SA3).

**Figure 4:**
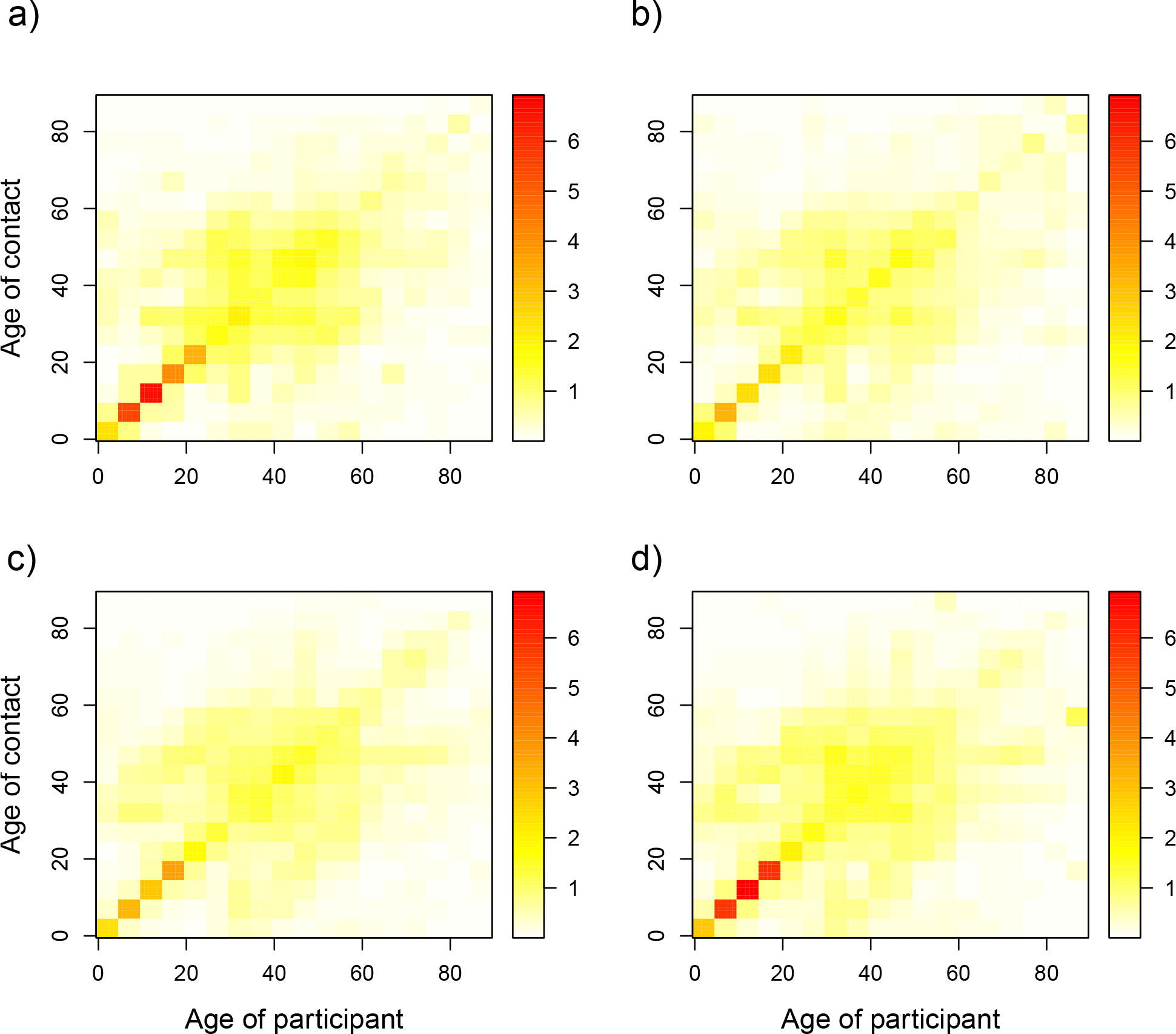
Estimated age- and gender-specific average number of contacts for the 2010-2011 survey. (a): male-male contacts, (b): male-female contacts, (c): female-male contacts and (d): female-female contacts. The color scale indicates the number of contacts from low (white) to high (red).

### School closure impact

We observed a significant difference in *R*_0_ between holiday and regular periods: the relative change in *R*_0_ equals 0.71 (BCI [0.60; 0.85]), or equivalently a 29% reduction in *R*_0_ for the holiday vs. the regular period. When comparing the relative change in *R*_0_ from a weekday to the weekend, a slightly higher reduction of 30% was observed. The difference in RI by age group is shown in Figure 5. The comparison of weekdays to weekends shows that the RI decreases significantly in the age group 0–15 years, while it is higher in the age group [60,65) and [70,75) on the weekend compared with the weekday. The RI also decreases from regular to holiday periods for the 3 age groups from 5 to 20 years, with the highest reduction observed in the age group 10–15 years. When comparing regular to holiday periods, we observe an increase in the RI for participants aged 65 to 80 years, though the RI variability for this age group is considerably high.

**Figure 5:**
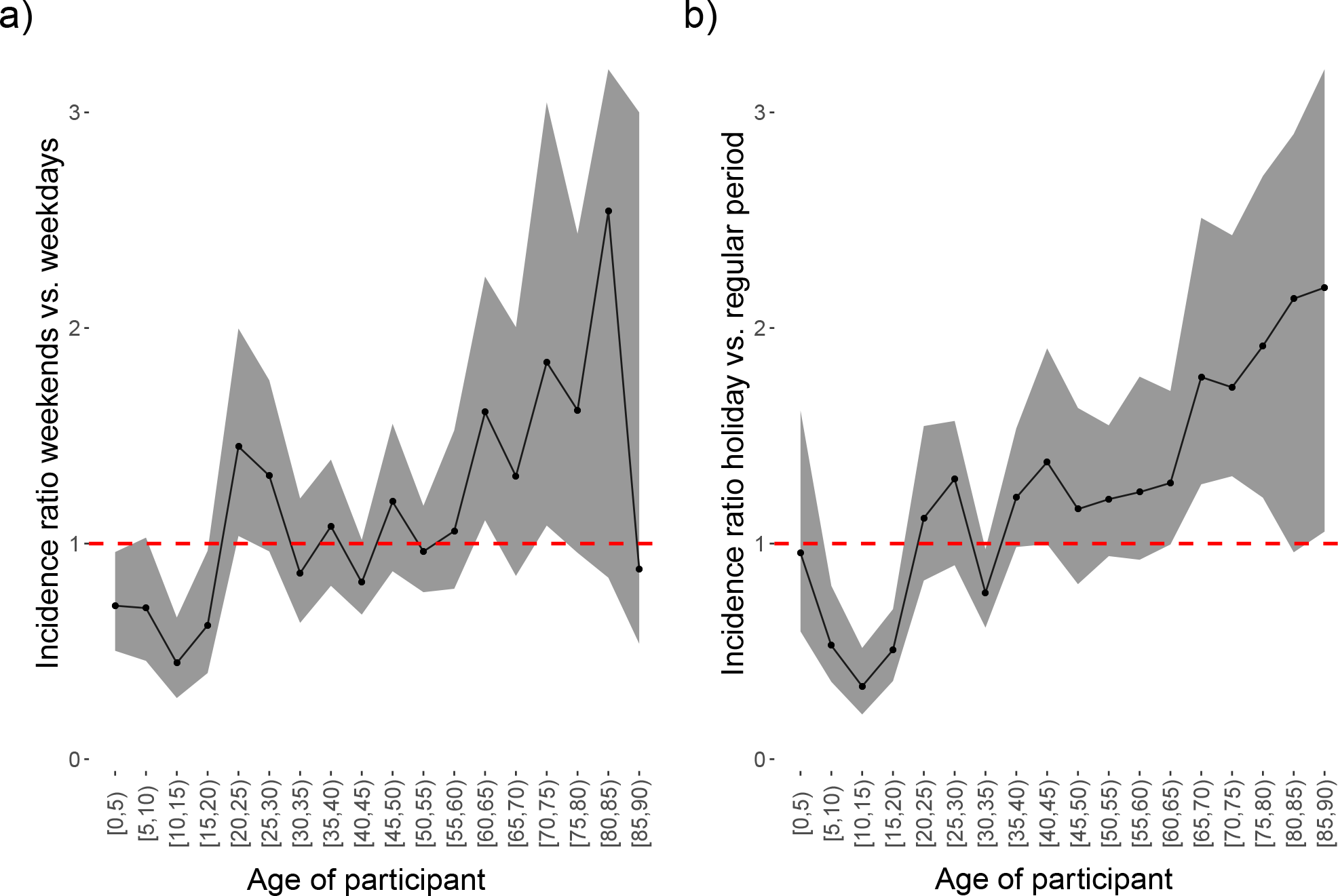
Ratio of relative incidences. (a): Ratio of relative incidences between weekends and weekdays. (b): Ratio of relative incidences between holidays and regular periods for the 2010-2011 survey. Shaded areas indicate 95% percentile bootstrap confidence intervals.

### Comparing the contact surveys in 2006 and 2010-2011

The age distribution of the Flemish population by age group does not change much in almost 5 years (SA3 Figure S4a). The distribution of participants in the survey 2010-2011 is closer to the Flemish population than that in the 2006 survey, especially for people older than 60 years. Participants aged between 0–20 years are over-sampled, while age groups 30–45 and 65–85 are under-sampled in the 2006 survey. The average number of contacts by age, adjusted for age, household size and types of days (weekdays/weekend days), in the two surveys is presented in SA3 Figure S4b. The largest difference in the number of survey participants is found for the age group 70–75.

### Degree distribution from aggregated data

The result of the random forest analysis is shown in Figure S5 SA3: gender yields the lowest mean decrease in accuracy, so it is removed. Significant predictors for both mean and over-dispersion parameters are further selected using the likelihood ratio test. Accordingly, the mean and over-dispersion regressions in the final model include household size and all interaction effects of three variables: age groups, microscopic time and macroscopic time.

The results of the GAMLSS model for the total number of contacts using the aggregated data from both surveys are shown in SA3 Table S4. After controlling for other factors, the household size is still significant, with a higher number of contacts recorded by participants living in a larger household. On regular weekdays, participants aged 60+ years reported the lowest number of contacts in both the 2006 and 2010-2011 surveys. During the same period, participants aged 45 –50 years had the highest number of contacts in the 2006 survey, while participants aged 30 –35 years had the highest number of contacts in the 2010-2011 survey. We observed a significant effect of microscopic time settings: in particular, participants aged 0–5 years reported a significantly lower number of contacts during weekends than regular weekdays, with RNC being 0.33 (CI [0.24; 0.45]) for the 2006 survey, while no significant differences in the number of contacts between during the regular weekday and the holiday weekdays was found for this age group. The main effect of macroscopic time (calendar year) and its interaction effects with age group and microscopic time settings are significant. On regular weekdays, participants aged 0–5 years old in the 2010-2011 survey reported a lower number of contacts than those in the 2006 survey: RNC is 0.64 (CI[0.46; 0.89]).

### Comparing the contact matrices for 2006 and 2010-2011

The difference between contact matrices estimated from the 2006 and 2010-2011 social contact surveys is negligible (Figure 6). Similar age and parent-child mixing patterns are observed for all contact matrices. The degree of assortativeness measured by Q and *I*^2^ indices (Table 2) are comparable in each microscopic time period, as evidenced by the overlapping BCIs. The relative incidences in 2006 and 2010-2011 (SA3 Figure S6) are also similar in both regular weekdays and holiday weekdays but are moderately dissimilar on the weekend, as the highest relative incidence is found in two different age classes (10-15 years and 20-25 years for 2006 and 2010-2011, respectively).

**Table 1:**
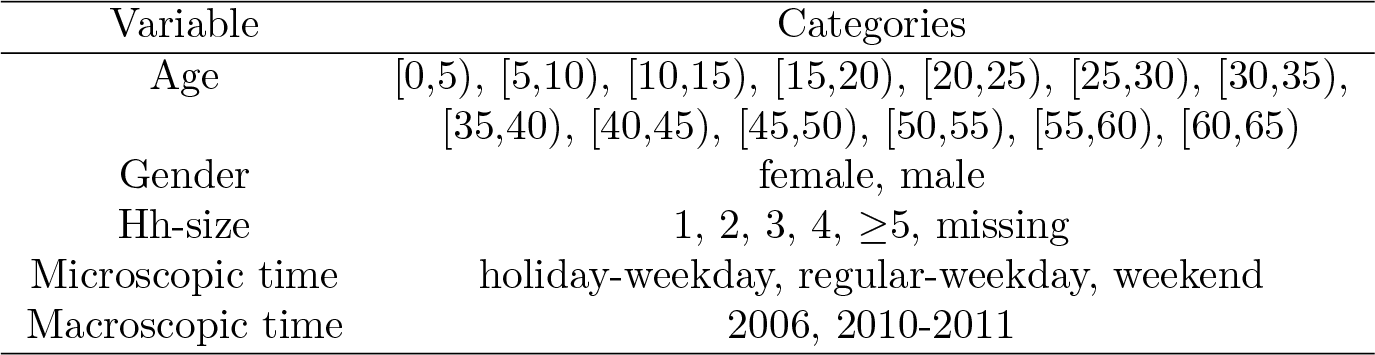
List of the common determinants selected from the two social contact surveys.

| Variable | Categories |
| --- | --- |
| Age | [0,5), [5,10), [10,15), [15,20), [20,25), [25,30), [30,35),<br>[35,40), [40,45), [45,50), [50,55), [55,60), [60,65) |
| Gender | female, male |
| Hh-size | 1, 2, 3, 4, $\geq 5$, missing |
| Microscopic time | holiday-weekday, regular-weekday, weekend |
| Macroscopic time | 2006, 2010-2011 |

**Table 2:**
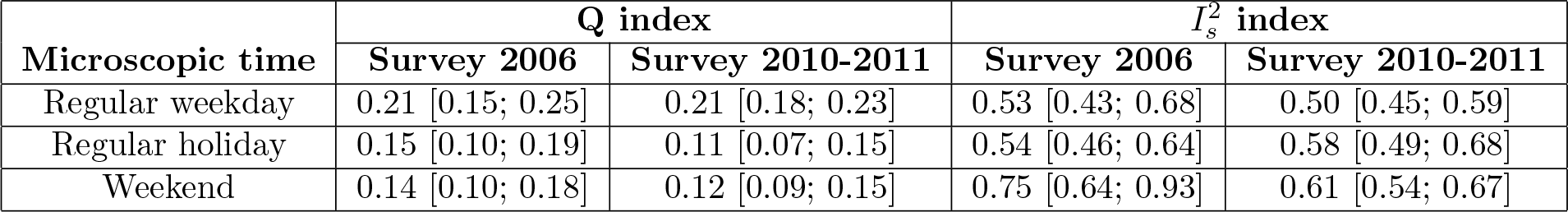
Assortativeness measures.

| Microscopic time | Q index | | $I_s^2$ index | |
| --- | --- | --- | --- | --- |
|  | Survey 2006 | Survey 2010-2011 | Survey 2006 | Survey 2010-2011 |
| Regular weekday | 0.21 [0.15; 0.25] | 0.21 [0.18; 0.23] | 0.53 [0.43; 0.68] | 0.50 [0.45; 0.59] |
| Regular holiday | 0.15 [0.10; 0.19] | 0.11 [0.07; 0.15] | 0.54 [0.46; 0.64] | 0.58 [0.49; 0.68] |
| Weekend | 0.14 [0.10; 0.18] | 0.12 [0.09; 0.15] | 0.75 [0.64; 0.93] | 0.61 [0.54; 0.67] |

**Figure 6:**
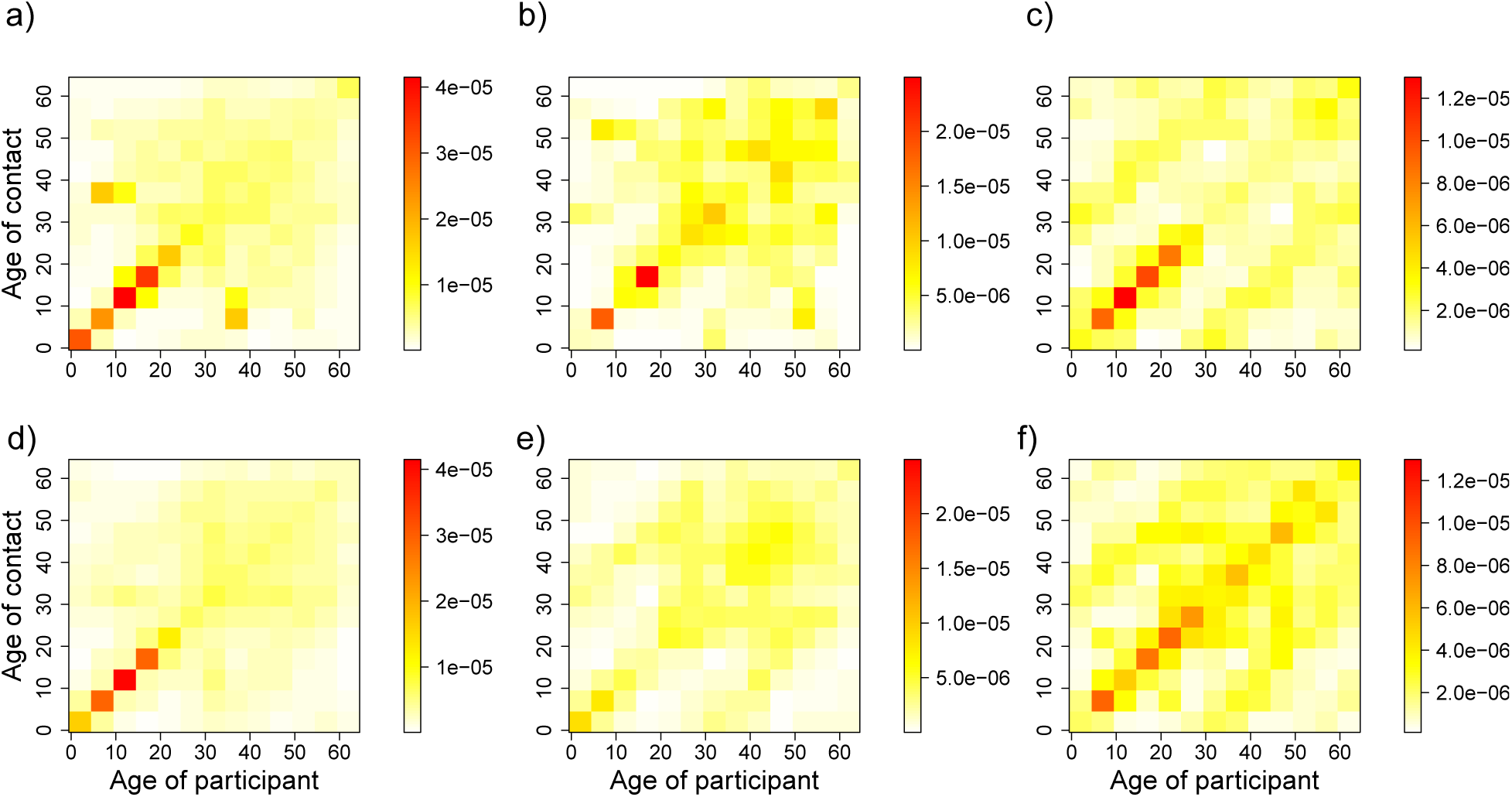
Age-specific contact rates. (a, d): Contact rates during regular weekdays, (b, e): Contact rates during holiday weekdays and (c, f): Contact rates during weekends. The 1st row represents the 2006 survey data, and the 2nd row represents the 2010-2011 survey data. The color scale indicates the contact rates from low (white) to high (red).

The relative change in *R*_0_, RRI and the ratio of transmission rates are used to further compare the epidemiological differences between contact matrices from the two surveys. The relative changes in *R*_0_ are presented in Figure 7 a). Based on the 95% BCI of relative changes in *R*_0_, significant changes between contact matrices of 2006 (numerator) and 2010-2011 (denominator) are only observed during the weekend, with an upper bound of the BCI close to 1. Figure 7 b) presents the changes in relative incidence between the 2006 and 2010-2011 social contact matrices stratified over 13 age groups and 3 microscopic time settings. Similar to the results of the relative change in *R*_0_, there was no evidence to support changes in RI over time, except for the age group 10-15 years during the weekend, where the RI was significantly higher in 2006 than in 2010-2011. The comparison of contact matrices based on the ratio of cell-wise contact rates in each microscopic time setting is provided in SA3 Figures S7, S8 and S9, for which the BCIs included a correction for multiple testings. We only found few significant differences in contact rates during holiday weekdays between 2006 and 2010-2011, mostly for participants aged 50+ years.

**Figure 7:**
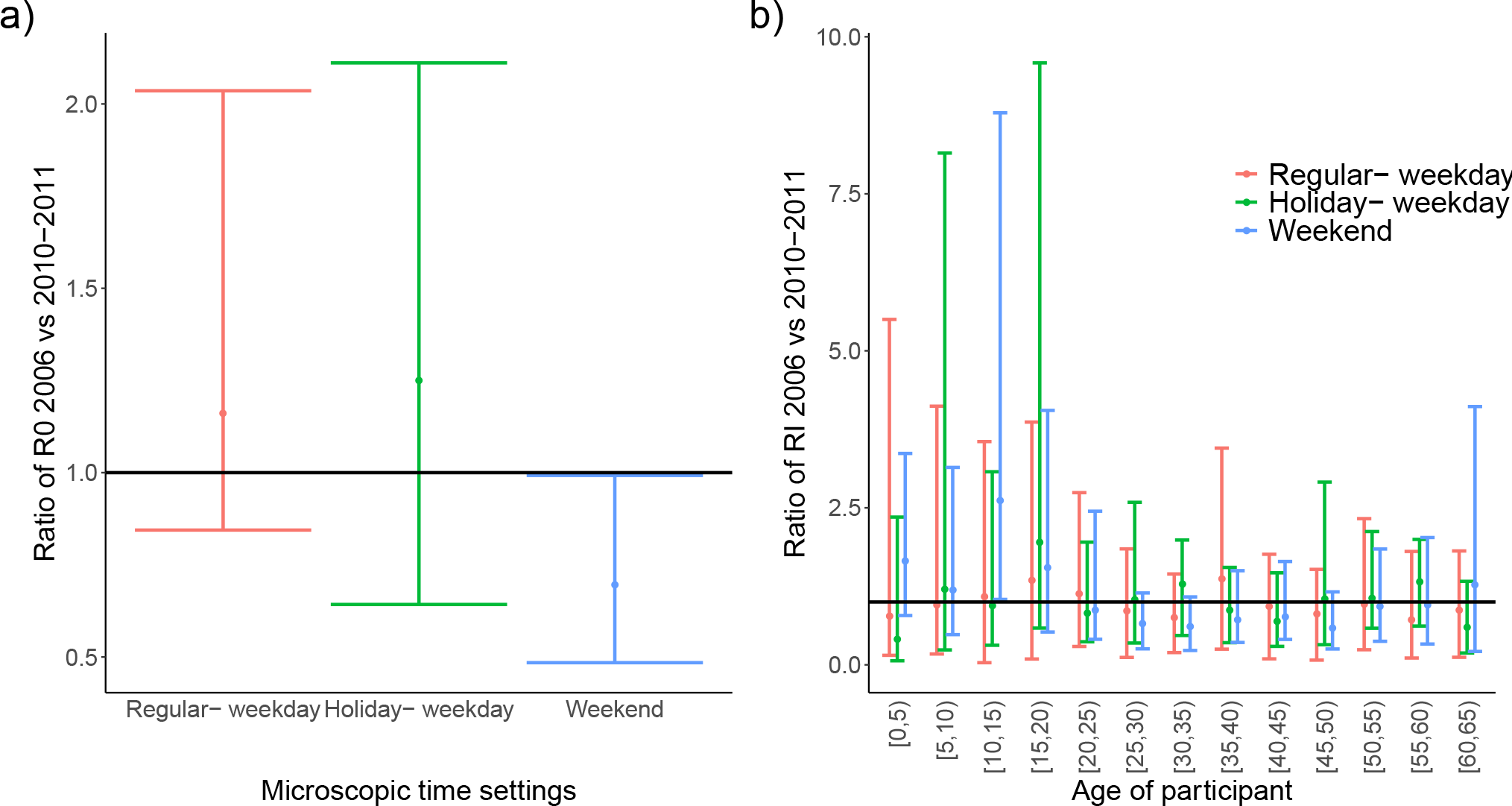
The ratio of basic reproduction numbers and of relative incidence by 5-year age groups. (a): The ratio of basic reproduction numbers and (b): the ratio of relative incidence by 5-year age groups for the 2006 and 2010-2011 social contact surveys in Flanders. The estimated relative changes in *R*_0_ and RRIs are shown by the dots with 95% BCI for each microscopic time setting. BCIs are obtained after Bonferroni correction with an overall significance level of 0.05. The horizontal line at 1 means no difference.

We compared the fit of the two contact matrices in 2006 and 2010-2011 to VZV serological data SA3 Figures S10. Although the contact matrix in 2006 produces a slightly better fit than the contact matrix in 2010-2011 (AIC: 1379 vs 1383), the observed prevalence and force of infection by age group between the two surveys almost completely overlap. We found no significant difference in *R*_0_ obtained from the contact matrices in 2006 and in 2010-2011 (10.7 BCI[6.23; 16.45] vs. 7.8 BCI [5.46; 7.51], respectively).

## Discussion and conclusions

Social contact surveys provide empirical data on populations’ mixing patterns that can inform mathematical models of infectious diseases. In Belgium, two large diary-based social contact surveys were conducted in 2006 and 2010-2011. In this work, we present the results of the latest survey, discussing the impact of microscopic time differences on mixing patterns and comparing the 2010-2011 data with the 2006 data. This approach allowed for assessing changes over macroscopic time differences, albeit in the limiting scenario of two surveys conducted only 4-5 years apart in a region where demography has remained fairly stable.

The association rules revealed that contacts of less than 15 minutes with non-household members usually do not involve skin-to-skin touching. This finding is in line with the results of the 2006 survey [28]. To investigate the contact profiles of participants, we performed clustering analysis. Our clustering results are comparable to the results in [3], in which a two-step clustering approach was applied to contact data from eight European countries. Specifically, we endorsed the “school profile”, “professional profile”, and “leisure profile” from [3], with more contacts during leisure activities during weekends.

Demographic factors, including age, household and province of residence, have significant effects on the number of contacts, as do the temporal factors, e.g., weekdays vs weekend days or regular terms vs holiday periods [9, 10, 16, 22, 40]. It is noted that the interaction between people aged 60+ years and young children/teenagers is significantly higher during holidays and weekends compared to regular-weekdays. For people living in an elderly/nursing home, however, almost no contacts with young children/teenagers are reported. Using public transportation is associated with a higher number of contacts in total. Our analysis also showed that those who reported to feel ill had fewer contacts than those who reported to be healthy [6, 10, 15, 16, 17]. This also holds for participants reporting health problems such as anxiety or those experiencing problems in daily activities.

There is evidence, at least among school-aged children, that contact patterns are assortative with respect to both age and gender. While an assortative mixing pattern with respect to age is still observed in adults, albeit with lower contact rates, an assortative mixing pattern with respect to gender disappears in people aged 30+ years. This analysis was also performed in [41], where a hierarchical Bayesian model was used to infer age-specific contact rates between genders. In contrast to [41], we did not find significant differences in infection risk between males and females. There are some reasons that may explain this difference. First, we aggregated the age of participants in 20 age classes instead of using continuous age, which can incur an inevitable loss of detail. Second, the dispersion parameter in our model was assumed to be age-dependent, while it was treated as a nuisance parameter in [41] to avoid computation challenges. In addition, we used diary weights in contact modelling to account for under-/over-sampling over the age of participants, while weights were not taken into account in the model of [41].

We found that the number of contacts was lower on weekends than on weekdays and during holidays compared to regular periods. We find a 30% (BCI:[17; 37%) reduction in *R*_0_ for weekends versus weekdays or a 29% (BCI:[14; 40%]) reduction in *R*_0_ for holidays versus regular periods. This result is consistent with the results of other studies [8, 19, 40, 42, 43]. However, computing the age-specific relative incidence showed that this reduction is due to the younger age classes, both during weekends and during holidays. Additionally, the age-specific relative incidence showed that during holidays, there is a more complex change than during regular weekends: while younger people have a lower relative incidence, people older than 60 years have an increased relative incidence during holidays.

Contact matrices were compared in different microscopic time settings, namely, regular weekdays, holiday weekdays and weekends, to obtain insights into possible changes in contact patterns between 2006 and 2010-2011 (Figure 6). Our result shows that irrespective of microscopic time settings, the contact patterns in 2006 and 2010-2011 follow the same trend of assortativeness. Furthermore, we observed pronounced inter-generational age mixing (the two sub-diagonals of the contact matrices), most likely indicating parent-child mixing patterns. This finding supports the evidence that households are central units in the epidemiology of airborne infections, e.g., influenza and SARS, because of the nature of the frequent and intimate contacts among household members. Children thus can have a bridging function, allowing for the spread of infection within households and to other households, from schools to workplaces or vice versa in a community [44]. The mixing patterns obtained from the contact matrices in 2006 and 2010-2011 in our study are in agreement with mixing patterns observed in similar studies [9, 10]. The relative incidences based on the 2006 and 2010-2011 data are quite similar between regular weekdays and holiday weekdays but are dissimilar on the weekend, as the highest relative incidence is found in two different age classes (10-15 years and 20-25 years for 2006 and 2010-2011, respectively).

In this study, we found that contact patterns remained fairly constant over 4-5 years. Additionally, within each microscopic time period, no substantial changes in the spread of infection, measured by the relative basic reproduction number and age-specific incidences, were observed (Figure 7). After taking into account multiple testings, the pair-wise comparison of contact rates over time present only few significant differences during holiday weekdays, mostly for people aged 50+ years. While the comparison of only two observational periods about five years apart can be considered a limitation, to the best of our knowledge, this is the first study, that investigates empirically whether contact rates remain stable, in the absence of major shocks to risk perception (as we expect to observe in the SARS-CoV-2 pandemic emergence year 2020) and demography. Hence our results suggest that stable social mixing patterns can be assumed over a time span of 5 years when no major shocks to risk perceptions or demography occur.

## Data Availability

The datasets analysed during the current study are available on Zenodo

https://doi.org/10.5281/zenodo.4059863

https://doi.org/10.5281/zenodo.4059825

## Declarations

### Availability of data and materials

The datasets analysed during the current study are available on Zenodo [45, 46].

### Competing interests

The authors declare that they have no competing interests.

### Author’s contributions

NH and PB designed and coordinated the survey. NH conceived the study and laid out a paper structure. LW and KVK performed data cleaning, TVH and YWK conducted the data analyses and drafted the manuscript in consultation with all the other authors. NH, PC and KVK made substantial revisions to the manuscript. All authors contributed to the final version of the manuscript. All authors approved the final manuscript and agreed with its submission to the journal.

### Ethics approval and consent to participate

Both datasets used for the current study are available online within the social contact data sharing initiative which is part of the ERC consolidator grant “TransMID” which received ethical approval from the Hasselt University Medical Ethical Committee (CME2016/618)

### Consent for publication

Not applicable.

### Funding

This work received funding from the European Research Council (ERC) under the European Union’s Horizon 2020 research and innovation programme (grant agreement 682540 — TransMID). LW gratefully acknowledge support from the Fonds voor Wetenschappelijk Onderzoek (FWO, postdoctoral fellowship 1234620N).

## Additional Files

**SA1 Descriptive data analysis of the 2010-2011 dataset**

**SA2 Data mining techniques**

**SA3 Supplementary results of the 2006 and 2010-2011 social contact survey data analyses**

## Notes

### Competing Interest Statement

The authors have declared no competing interest.

### Author Declarations

The Hasselt480University Medical Ethical Committee (CME2016/618)

